# Genital Inflammatory Responses in Women Living with HIV Randomized to Copper or Levonorgestrel Intrauterine Contraceptives: A secondary analysis of a randomized trial

**DOI:** 10.64898/2026.05.24.26353969

**Authors:** Anna-Ursula Happel, Jo-Ann S. Passmore, Musalula Sinkala, Shameem Z. Jaumdally, Hoyam Gamieldien, Nai-Chung Hu, Nontokozo Langwenya, Heidi E. Jones, Donald R. Hoover, Landon Myer, Catherine S. Todd

## Abstract

**Background:** Intrauterine contraceptives (IUCs) are effective, but effects on genital inflammation among women living with HIV (WLHIV) by antiretroviral therapy (ART) use are unclear. We evaluated the longitudinal effects of copper IUC (CLIUC) and the levonorgestrel intrauterine system (LNGLIUS) on cervicovaginal cytokine profiles in a secondary analysis of a randomized trial (NCT01721798, 2013–2016).

**Methods:** Cervicovaginal secretions were collected from 100 WLHIV (nonLART usersL=L48; ART usersL=L52) randomized 1:1 to ClZIUC or LNGlZIUS. TwentyLeight cytokines were measured prior to insertion and 3- and 6-months postLinsertion. Cytokine concentrations at each followLup visit were compared with baseline, using participant fixedLeffects models stratified by ART status.

**Results:** At enrollment, nonLART users had higher average concentrations of most cytokines (21/28) than women using ART. Among non-ART users, IUC use was not associated with cytokine increases; only MCPL1 increased significantly at 3 months among CLIUC users (log_10_ geometric mean ratio 0.77, 95%CI 0.38–1.17), while none increased with LNG-IUS use. Among ART users, C-IUC insertion resulted in broad and sustained cytokine increases at both 3 (16/28) and 6 months (15/28). At month 3, the largest increases in log_10_ geometric mean were observed for IL-6 (1.04, 0.72-1.36), RANTES (0.97, 0.54-1.40), MCP-1 (0.71, 0.46-0.96), MIP-1α (0.66, 0.37-0.94), and G-CSF (0.63, 0.36-0.89), which was maintained until month 6. Cytokine changes following LNGLIUS insertion were minimal (IL-5, month 3).

**Conclusions:** Among ART users, CLIUC is associated with increases in cervicovaginal cytokines, across functional classes. This supports LNGLIUS as a less inflammatory option for WLHIV to minimize genital immune activation.

## Introduction

Safe, effective and acceptable contraception is essential for voluntary, rights-based family planning programs, including equitable access for women living with HIV (WLHIV) [1]. Despite expanded antiretroviral therapy (ART) access and use, WLHIV continue to experience high rates of unmet need for modern contraceptives [2,3] and mistimed or unwanted pregnancies [4,5], driven in part by ongoing concerns about contraceptive safety and interactions with ART [6–8]. Ensuring both clinical safety and biologic compatibility of contraceptive methods with ART remain essential to expanding informed choices for WLHIV.

Use of long-acting reversible contraceptives (LARCs) has increased substantially in sub-Saharan Africa, with marked growth in uptake of progestin-only implants and copper intrauterine contraceptives (C-IUC) over the past decade [9]. IUCs are among the most effective available LARC method and have demonstrated clinical safety in WLHIV, including no significant increase in pelvic inflammatory disease risk, HIV disease progression, or genital HIV shedding [10–12]. The two IUCs most studied in this population, the non-hormonal C-IUC and the levonorgestrel intrauterine system (LNG-IUS), are increasingly available in low- and middle-income countries with high HIV prevalence [13].

In women without HIV (WWOH), IUC insertion induces well-described morphological, biochemical and inflammatory responses within the reproductive tract, including leukocyte infiltration and upregulation of proLinflammatory cytokines and chemokines. Although initially thought to be confined to the uterus and endometrium [12,13], these immune changes are now recognized to extend to the lower genital tract [16,17]. Genital inflammation is a key driver of HIV shedding in genital secretions, even among women using effective ART [16,17],. Whether and how IUC-associated immune responses differ among WLHIV by ART use, and if different IUC types cause different responses, is unknown.

Here, we performed a secondary analysis of a randomized clinical trial in which WLHIV were randomized to CLIUC or LNGLIUS. We wanted to understand first, whether there were differences in cytokine profiles by ART use prior to IUC insertion; second, how cytokine profiles changed over time within groups of IUC users (stratified by ART use), and third whether cytokine trajectories post IUC insertion differed by IUC (stratified by ART use). Our hypothesis was that C-IUC insertion would be associated with greater increases in proLinflammatory cervicovaginal cytokines, as seen in WWOH [17,20,21], compared to LNG-IUS, in ART users and non-ART users.

## Methods

This was secondary analysis of a double-masked randomized controlled trial with one-to-one allocation to the C-IUC (SMB Copper T-380A IUD; SMB Corporation of India, Mumbai, India, or Nova-T Copper T-380 IUD; Bayer Pharmaceuticals, Germany) and the LNG-IUS (Mirena; Bayer Health Care Pharmaceuticals, Montville, NJ, stratified by ART use, age and recent injectable progestin contraceptive exposure at enrollment), among WLHIV in Gugulethu, Cape Town, South Africa (clinicaltrials.gov NCT01721798, https://clinicaltrials.gov/study/NCT01721798, registered on 18 Oct 2012) [22]. Women were recruited from surrounding health facilities, community events promoting reproductive health, and radio and newspaper advertisements between October 2013 to December 2016, and clinical activities closed in July 2018.

### Ethics

Ethical approval was obtained from the human research ethics committees of the University of Cape Town, South Africa (283/2012) and FHI 360, United States (398733-61). All women provided written informed consent prior to enrollment.

### Cohort

Comprehensive details on the protocol can be found in the primary paper [22]. Briefly, eligible women were 18-40 years old, had a confirmed HIV diagnosis, were not pregnant or wanting to become pregnant in the next 30 months, at least 6 months post-partum, not planning relocation within 30 months and were without any condition contraindicating IUC use. Eligible WLHIV were either 1) virally-supressed (plasma viral load [pVL] < 1,000 copies/mL) on ART, or 2) were ART-ineligible at enrollment by CD4 lymphocyte count per local guidelines at the time [22]. At screening, women were tested for reproductive tract infections (RTIs) *Neisseria gonorrhea* [NG], *Chlamydia trachomatis* [CT], *Trichomonas vaginalis* [TV], *Treponema pallidum* [TP], and sialidase-positive bacterial vaginosis [BV] and treated based on reactive tests. Women testing negative at screening or between two and four weeks post-treatment for any RTIs were eligible to enroll. For this secondary analysis, we selected the first consecutively enrolled women in each randomization and ART stratum (CLIUC/ART users, CLIUC/nonLART users, LNGLIUS/ART users, LNGLIUS/nonLART users) who had completed 6-month cervicovaginal sampling.

### Randomization

In the parent randomized trial, participants were allocated 1:1 to CLIUC or LNGLIUS using permuted blocks, stratified by ART status at enrolment, age category (18-23, 24-31, 32-40), and recent (<=6 months) injectable contraceptive exposure. An independent data manager generated the sequence and prepared stratumLspecific, sequentially numbered, opaque sealed envelopes; site staff opened the next envelope at enrolment to assign group. Study nurses performing IUC insertions and a site manager were unmasked; all investigators, laboratory personnel, and clinical outcome assessors remained masked. Participants were masked during followLup and unmasked at the exit visit.

### Visit procedures

At enrollment, participants completed a questionnaire on sociodemographics, sexual history, menstrual bleeding, and pelvic symptoms, and were randomized to C-IUC or LNG-IUS insertion. At all three visits (pre-IUC insertion [baseline], 3 and 6 months post-IUC), participants provided urine specimens for pregnancy testing, blood for pVL testing, and inserted menstrual cups (MC, Instead Softcup^TM^, Evofem Inc., San Diego, CA, USA) to collect cervico-vaginal secretions for genital viral load (gVL) [22] and cytokine testing [23]. In the laboratory, MCs were weighed in Falcon tubes and the volume of secreted fluid measured. Assuming 1g=1mL genital fluid, MC secretions were diluted at a 9:1 ratio with phosphate buffered saline (PBS), then stored at -80°C [24]. Visits were scheduled to avoid menstrual bleeding, and participants were asked to avoid vaginal intercourse or douching 48 hours prior to the visit.

### Laboratory testing

HIV RNA concentrations in plasma and cervicovaginal secretions were measured at the South African National Health Laboratory Service (NHLS). Genital tract specimens were tested for NG and CT (NG/CT Xpert®, Cepheid Diagnostics, Sunnyvale, California, USA), TV (OSOM® Trichomonas) and sialidase+BV (OSOM® BV Blue, Sekisui Diagnostics, Lexington, Massachusetts, USA) at screening and the 3 and 6 month visits. Plasma was tested for TP (Alere® Determine® Syphilis, Alere Diagnostics, San Diego, California, USA) [22].

### Measurement of genital tract cytokines by Luminex

Concentrations of 27 cytokines were measured using a Bio-Plex Pro Human Cytokine 27-plex Assay (Bio-Rad Laboratories Inc., USA), and IL-1α was measured using a single bead array [23]. Cytokines were functionally grouped into inflammatory (IL-1α, IL-1β, IL-6, IL-12p70, TNF-α); chemotactic (eotaxin, IL-8, interferon gamma-induced protein [IP]-10, MCP-1, macrophage inflammatory protein [MIP]-1α, MIP-1β, RANTES); adaptive (IFN-γ, IL-2, IL-4, IL-5, IL-13, IL-15, IL-17); hematopoietic (growth factors; fibroblast growth factor [FGF]-basic, granulocyte colony-stimulating factor [G-CSF], GM-CSF, IL-7, IL-9, platelet-derived growth factor [PDGF]-bb, vascular endothelial growth factor [VEGF]); and anti-inflammatory cytokines (IL-10, IL-1 receptor antagonist [RA]). Plates were analyzed using a Bio-Plex Suspension Array Reader (Bio-Rad Laboratories Inc., USA) and Bio-Plex manager software (version 4). Cytokine concentrations below the lower limit of detection were reported as the mid-point between zero and the lowest concentration measured for a given cytokine. A panel of nine repeat samples were included on each Luminex plate to determine intra-assay and inter-assay correlation coefficients [23].

### Outcomes

Detectable gVL was the primary trial outcome, while plasma VL (pVL), occurrence of adverse events, and IUC acceptability were secondary outcomes [22]. The outcomes of interest of this secondary analysis were: (i) changes in cervicovaginal cytokine concentrations from baseline to 3 and 6 months post-insertion, and (ii) longitudinal cytokine trajectories over the 6 month followLup period, evaluated within each IUC and ART stratum (CLIUC/ART users, CLIUC/nonLART users, LNGLIUS/ART users, LNGLIUS/nonLART users). Exploratory analyses included identifying cytokine markers that most effectively distinguished between CLIUC/ART users and LNGLIUS/ART users.

### Sample power considerations

Using ILL6 foldLchange data (median 10.1Lfold increase; interquartile range (IQR) 1.5–83.6 from baseline to 6 months) reported in the ECHO trial that randomized WWOH to C-IUC [17], and assuming a logLnormal distribution, the corresponding standard deviation (SD) on the logLscale was estimated as 2.98. With 25 women per stratum, this secondary analysis had approximately 83% power at α = 0.05 to detect a change in ILL6 over 6 months equivalent to the previously observed effect size. Based on actual sample sizes (22–29 per stratum), power ranged from 78 to 86%.

### Statistical Analysis

Analyses were performed using R (v4.2.2) [25]. Participant characteristics were summarized using means (± standard deviations), medians (with interquartile ranges), and proportions at enrolment by randomization group. Missing data were left missing, and analyses used all available data points. We addressed three interrelated research questions: (1) whether baseline cytokine levels differed by ART use prior to IUC insertion; (2) whether there were within-participant changes in cytokine concentrations following LNG-IUS and C-IUC insertion at months 3 and 6 relative to baseline; and (3) whether longitudinal cytokine trajectories over the 6-month follow-up period differed by IUC type. To address the first question, we compared baseline (pre-insertion) cytokine concentrations between ART groups (ART users vs. non-users) using the Wilcoxon rank-sum test, and p values were adjusted for multiple comparisons using the Bonferroni correction across all cytokines. To address the second and third question, we fitted linear regression models with participant-specific fixed intercepts to account for between-participant heterogeneity, thereby controlling for all time-invariant participant characteristics, including baseline cytokine levels. Analyses were conducted separately by ART status (ART vs non ART). For each cytokine, raw concentrations were log_10_ transformed to improve normality. Time was modeled as a categorical variable with baseline as the reference category, allowing estimation of changes at month 3 relative to baseline and at month 6 relative baseline. IUC group was included as a categorical factor (LNGLIUS as the reference group vs. CLIUC). Because IUC assignment was time-invariant within a participant (all participants remained on their assigned IUC type), its main effect is absorbed by the participant fixed effect and was therefore not separately identified. However, we included an interaction term between time and IUC assignment to assess whether post-insertion trajectories differed by IUC type. The model was specified as: log _10_(cytokine_it_)= a_i_ + p_time_ + p_timeXIUD_ + c_it_, where a_i_ represents a participantLspecific fixed effect and c_it_is the error term. To address the second question, we used linear combinations of fitted model coefficients (contrasts) to estimate changes in cytokine concentrations from baseline to month 3 and from baseline to month 6 within each IUC group, expressed as geometric mean ratios (GMRs). To address the third question, for each cytokine, joint F-tests were conducted to evaluate (i) the overall effect of time and (ii) the time × IUC interaction, testing whether longitudinal cytokine changes differed by IUC type. All hypothesis tests were derived from the fitted fixed-effects models. Both raw and Benjamini-Hochberg-adjusted p-values are reported. The false discovery rate was controlled within each ART stratum, with the family defined as all cytokines and post-insertion time-point contrasts evaluated.

Finally, we used unsupervised hierarchical clustering with a cosine distance metric using complete linkage to compare the patterns of genital cytokines levels between the ART and IUC groups at baseline, Month 3 and Month 3. Given that cytokine changes were mainly occurring among ART users only, we applied Partial Least Squares Discriminant Analysis (PLSLDA) [26] to compare baseline to month 6 changes in cytokine concentrations between CLIUC and LNGLIUS users in this group. The IUC group served as the classification outcome, and model components were selected using crossLvalidation to minimize classification error. VariableLimportanceLinLprojection (VIP) scores were generated from the final model to identify cytokines contributing most strongly to group separation; cytokines with VIP ≥1 were considered influential.

## Results

The 100 WLHIV in this analysis were enrolled and provided samples between November, 2013 and February, 2017 for non-ART and between April, 2015 and June, 2017 for ART-using participants (Supplementary Figure 1). At enrolment, these women had a median time since HIV diagnosis of seven years (IQR 4-9), with most reporting having a current male partner (92.0%, 92/100) (Table 1). Few women had bacterial vaginosis (15.0%; 15/100), *N. gonnorhoea* (6.0%; 6/100), *C. trachomatis* (6.0%; 6/100), or *T. vaginalis* (9.0%; 9/100); prevalences did not differ by randomization group. At enrollment, 52 WLHIV reported ART use. ART users generally were virally suppressed (pVL <40 copies/ml; 84.6%, 44/52). Among those with detectable pVL, values were low, with a median pVL of 2.49 log10 copies/mL (IQR 2.14–3.28 log10). Genital VL was undetectable (defined as <40 copies/ml in menstrual cup samples) in 92.3% (48/52) of ART users. Non-ART using WLHIV (n=48) had median pVLs of 3.77 log10 copies/mL (IQR 3.17–4.29 log10); and 60.4% (29/48) had detectable gVL, with a median gVL of 3.55 log10 copies/mL (IQR 3.04–4.11 log10). None of the 48 nonLART users initiated ART prior to month 6, the final time point included in this analysis.

**Table 1.** Cohort characteristics.

|  | All | C-IUC | LNG-IUS |
| --- | --- | --- | --- |
| N | 100 | 45 | 55 |
| Age, years (median; IQR) | 30 (27-33) | 30 (27-33) | 30 (28-34) |
| Years since HIV diagnosis (median; IQR) | 7 (4-9) | 7 (4-10) | 6 (4-9) |
| ART status |  |  |  |
| Non-ART (n; %) | 48 (48.0) | 22 (48.9) | 26 (47.3) |
| ART (n; %) | 52 (52.0) | 23 (51.1) | 29 (52.7) |
| Current partner (n/N; %) | 92 (92.0) | 40 (88.9) | 52 (94.4) |
| Reproductive tract infections at screening (n; %)<sup>@</sup> |  |  |  |
| *C. trachomatis* | 6 (6.0) | 2 (4.4) | 4 (7.2) |
| *N. gonorrhoea* | 6 (6.0) | 2 (4.4) | 4 (7.2) |
| *T. vaginalis* | 9 (9.0) | 3 (6.7) | 6 (10.9) |
| HSV-2 [shedding] | 6 (6.3)<sup>a</sup> | 3 (7.0)<sup>a</sup> | 3 (5.8)<sup>a</sup> |
| HSV-2 [IgG] | 88 (94.6)<sup>b</sup> | 38 (92.7)<sup>b</sup> | 50 (96.2)<sup>b</sup> |
| Bacterial vaginosis<sup>#</sup> | 15 (15.0) | 4 (8.9) | 11 (20.0) |

<sup>@</sup>Women were treated for RTIs prior to enrollment. <sup>#</sup>Determined by OSOM® BV Blue. <sup>a</sup>missing data n=5 (C-IUC n=2, LNG-IUS n=3). <sup>b</sup>missing data n=7 (C-IUC n=4, LNG-IUS n=3). There were no statistical differences at p = 0.05

### Baseline genital tract cytokine profiles stratified by ART use

At enrollment, cervicovaginal concentrations of 21/28 cytokines (75.0%) differed significantly by ART use, after adjusting for multiple comparisons. Of those, 14 cytokines (IL-1α, IL-1β, Il-6, IL-8, IP-10, MIP-1α, MIP-1β, RANTES, IL-7, PDGF-bb, VEGF, IFN-γ, IL-13 and IL-10) were significantly higher in non-ART compared to ART-using WLHIV, while seven (Eotaxin, FGF basic, GM-CSF, IL-9, IL-2, Il-4 and IL1-RA) were significantly higher in ART compared to non-ART-using WLHIV, across all functional cytokine classes (Table 2). Similar findings were noted when stratifying participants based on the detectability of gVL at baseline (Supplementary Table 1). Therefore, all subsequent analyses were conducted stratified by ART status at baseline.

**Table 2.** Genital tract cytokines at enrollment in non-ART and ART-using WLHIV.

| Class | Cytokine | Non-ART (n=48)<br>Median [IQR] | ART (n=52)<br>Median [IQR] | Adjusted<br>p-value<sup>#</sup> |
| --- | --- | --- | --- | --- |
| Inflammatory | IL-1α | 822.82 [287.14, 1363.92] | 415.95 [163.48, 865.46] | **0.030** |
| | IL-1β | 913.72 [189.74, 1543.81] | 106.06 [36.15, 409.02] | **0.002** |
|  | IL-6 | 50.12 [13.72, 121.63] | 10.57 [5.08, 24.51] | **0.002** |
|  | IL-12(p70) | 77.42 [60.00, 97.72] | 67.11 [41.42, 93.66] | 0.199 |
| | TNF-α | 38.21 [13.43, 71.72] | 39.15 [27.56, 63.35] | 0.330 |
| Chemokines | Eotaxin | 4.17 [0.98, 10.76] | 7.83 [6.19, 10.06] | **0.005** |
|  | IL-8 | 1664.18 [741.76, 2998.40] | 838.31 [356.40, 1901.46] | **0.060** |
|  | IP-10 | 2183.70 [284.59, 5877.95] | 355.36 [98.38, 1314.94] | **0.010** |
|  | MCP-1 | 21.72 [14.26, 57.84] | 36.72 [20.75, 54.60] | 0.120 |
| | MIP-1α | 3.13 [1.90, 4.93] | 1.22 [0.81, 2.30] | **0.002** |
| | MIP-1β | 74.25 [43.66, 213.24] | 24.86 [12.24, 57.22] | **0.002** |
|  | RANTES | 35.26 [13.90, 86.71] | 3.72 [1.80, 13.84] | **0.002** |
| Growth factors | FGF basic | 19.39 [14.33, 25.44] | 39.24 [33.72, 44.05] | **0.002** |
|  | G-CSF | 1964.00 [511.40-4951.00] | 804.30 [334.00-2876.00] | 0.060 |
|  | GM-CSF | 66.29 [51.91, 82.98] | 108.60 [76.05, 130.94] | **0.002** |
|  | IL-7 | 4.89 [3.66, 7.99] | 3.71 [2.41, 5.50] | **0.005** |
|  | IL-9 | 4.90 [3.30, 6.65] | 18.28 [13.06, 25.15] | **0.002** |
|  | PDGF-bb | 29.13 [17.45, 41.55] | 13.84 [9.98, 19.89] | **0.002** |
|  | VEGF | 2423.33 [1326.24, 3698.32] | 1012.63 [542.25, 1745.71] | **0.002** |
| Adaptive | IFN-γ | 49.37 [29.44, 77.34] | 23.83 [17.72, 30.37] | **0.002** |
|  | IL-2 | 0.02 [0.02, 2.77] | 2.42 [0.40, 3.68] | **0.002** |
|  | IL-4 | 0.84 [0.55, 1.35] | 1.15 [0.86, 1.46] | **0.008** |
|  | IL-5 | 0.52 [0.08, 3.42] | 1.04 [0.04, 3.19] | 0.930 |
|  | IL-13 | 2.72 [1.91, 3.55] | 1.00 [0.71, 1.38] | **0.002** |
|  | IL-15 | 2.59 [0.01, 6.29] | 2.69 [0.14, 9.40] | 0.088 |
|  | IL-17 | 26.48 [13.86, 37.54] | 19.59 [14.59, 27.72] | 0.504 |
| Anti-inflammatory | IL-10 | 16.31 [14.35, 20.37] | 10.72 [8.04, 13.98] | **0.002** |
|  | IL-1RA | 5.8x10<sup>4</sup> [0.9, 10.0x10<sup>4</sup>] | 1088.9x10<sup>4</sup> [227.2, 1088.9x10<sup>4</sup>] | **0.002** |

<sup>#</sup>P-values were adjusted for multiple comparisons using Bonferroni across all ART vs. non ART using comparisons.

### Changes in cervicovaginal cytokine concentrations following IUC insertion

#### VisitLspecific cytokine changes

Demographics, prevalence of STIs at screening, and cytokine concentrations at baseline were comparable between ART non-users (Supplementary Table 2) and ART users (Supplementary Table 3) randomized to C-IUC versus LNG-IUS. Among non-ART users, C-IUC insertion resulted in an increase of five cytokines (MCP-1, MIP-1α, IL-15, IL-13 and G-CSF, p<0.05) at 3 months post-insertion compared to baseline (Figure 1), but only MCP-1 remained statistically significant after multiple comparison adjustment (log_10_ GMR 0.77, 95% CI 0.38 – 1.17, adj. p = 0.02). At 6 months, MCP-1, MIP-1α, IL-15 and G-CSF were still increased compared to baseline, in addition to IL-5, IL-10, and IL-17, although none were statistically significant after multiple comparison adjustment. Among non-ART LNG-IUS users, changes in cytokine profiles were more variable, with an increase of MCP-1 and decrease of IP-10 at 3 months post-insertion compared to baseline, and increased IL-2, MIP-1α and MIP-1β, IL-15, MCP-1, IL-6 and G-CSF at month 8 compared to baseline. However, none was statistically significant multiple comparison adjustment (Figure 1, Supplementary Table 4). Overall, these findings suggest that most cervicovaginal cytokine concentrations at 3 and 6 months did not differ from baseline following CuLIUC or LNGLIUS insertion among non-ART users.

**Figure 1.**
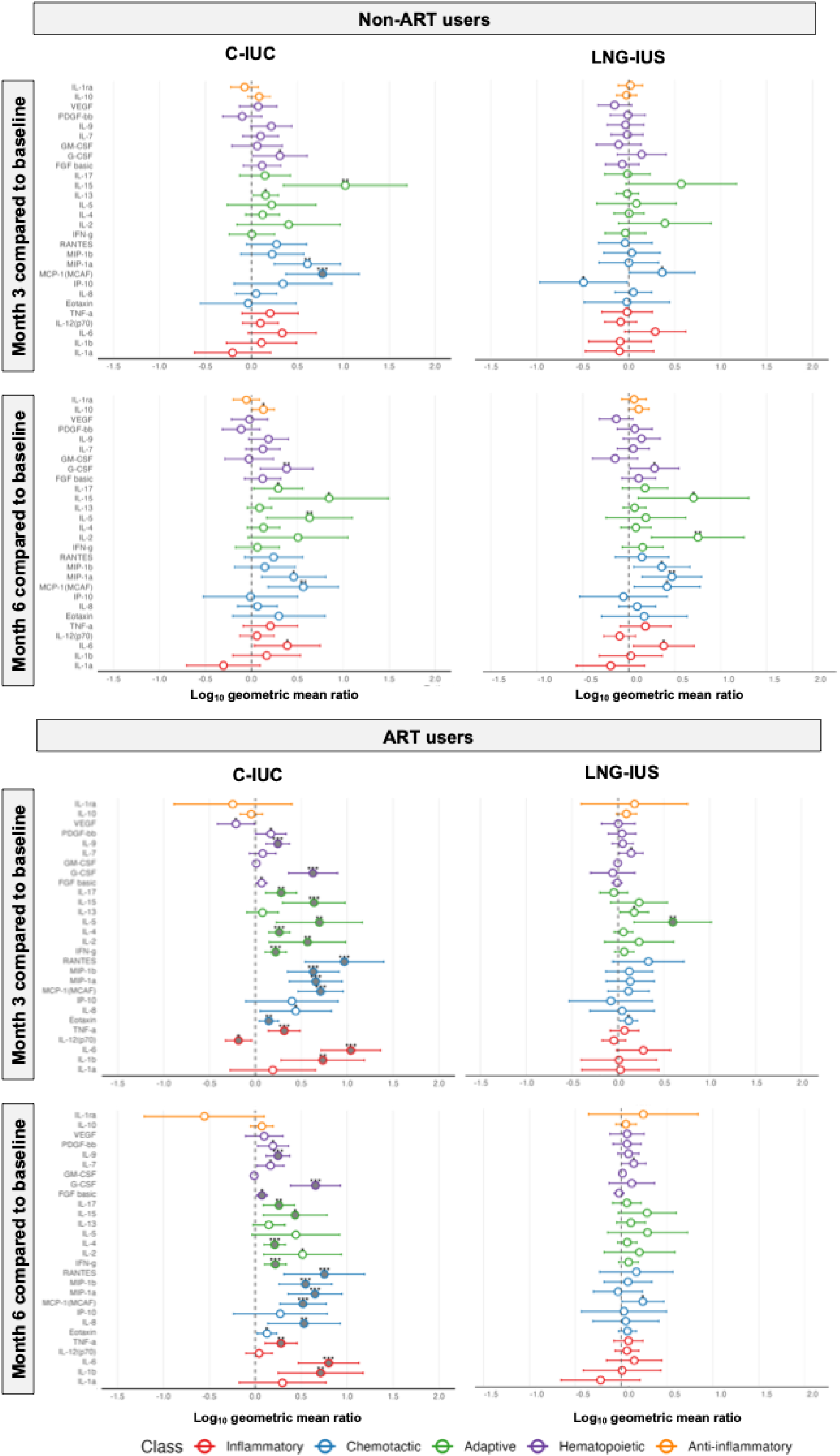
Log_10_ fold-change of genital tract cytokine concentrations at months 3 and 6 relative to baseline in WLHIV using C-IUC and LNG-IUS, faceted by ART use. Forest plots display geometric mean ratios (GMRs, circles) and 95% confidence intervals (horizontal bars) from participant fixedLJeffects models comparing cytokine concentrations at Month 3 vs. baseline (top) and Month 6 vs. baseline (bottom). GMRs are presented on the original log_10_ scale, with positive values indicating higher cytokine concentrations relative to baseline, and negative values indicating lower concentrations. Results are faceted by ART status (NonLJART = left panel, ART users = righ panel) and contraceptive type (CLJIUC and LNGLJIUS). Cytokines were grouped by functional class: inflammatory (green), growth factors (yellow), chemokines (purple), anti-inflammatory cytokines (red) and adaptive cytokines (blue). Open circles represent the raw effect estimate. Stars above circles correspond to unadjusted pLJvalues (*p < 0.05; **p < 0.01; **p < 0.001) for comparisons of concentrations at months 3 and 6 relative to baseline. Filled circles indicate that adjusted p < 0.05. P-values were adjusted for multiple comparisons using the Benjamini–Hochberg false discovery rate procedure within ART strata, across all cytokines and timeLJpoint contrasts evaluated.

Women using ART had significantly lower levels of most cervicovaginal cytokines at baseline than non-ART using women (Table 2). Among ART users, CLIUC insertion was associated with widespread (unadjusted p<0.05) cytokine increases at both 3 months (19/28 cytokines) and 6 months (19/28 cytokines) postLinsertion relative to baseline, across all functional classes. After Benjamini-Hochberg multipleLcomparison adjustment, 16 of these increases at 3 months remained significant (16/28: IL-6, MCP-1, G-CSF, IL-4, MIP-1a, RANTES, MIP-1b, IL-9, IFN-g, IL-15, TNF-a, IL-17, IL-1b, IL-5, Eotaxin and IL-2) and 15 at 6 months (15/28: IL-6, G-CSF, MIP-1a, MCP-1, IL-9, MIP-1b, IFN-g, IL-4, RANTES, TNF-a, IL-1b, IL-17, IL-8, FGF basic and IL-15) relative to baseline (Figure 1, Supplementary Table 4). In contrast, IL-12-p70 and VEGF were significantly reduced after 3 months but not at 6 months following C-IUC insertion in ART users. Changes in cervicovaginal cytokines in the ART-using LNG-IUS group were less pronounced. At 3 months post-insertion, four cytokines were significantly increased (IL-5, Eotaxin, IL-13 and IL-7, of which only IL-5 remained significant after multiple comparison adjustment) and only MCP-1 and IL-7 were increased at 6 months post-insertion relative to baseline prior to multiple comparison adjustment (Figure 1, Supplementary Table 4).

#### Longitudinal patterns

To assess whether patterns of cytokine change across study visits differed by IUC type, we evaluated the overall effect of time and timeL×LIUC group interaction (Figure 2; Supplementary Table 5).

**Figure 2.**
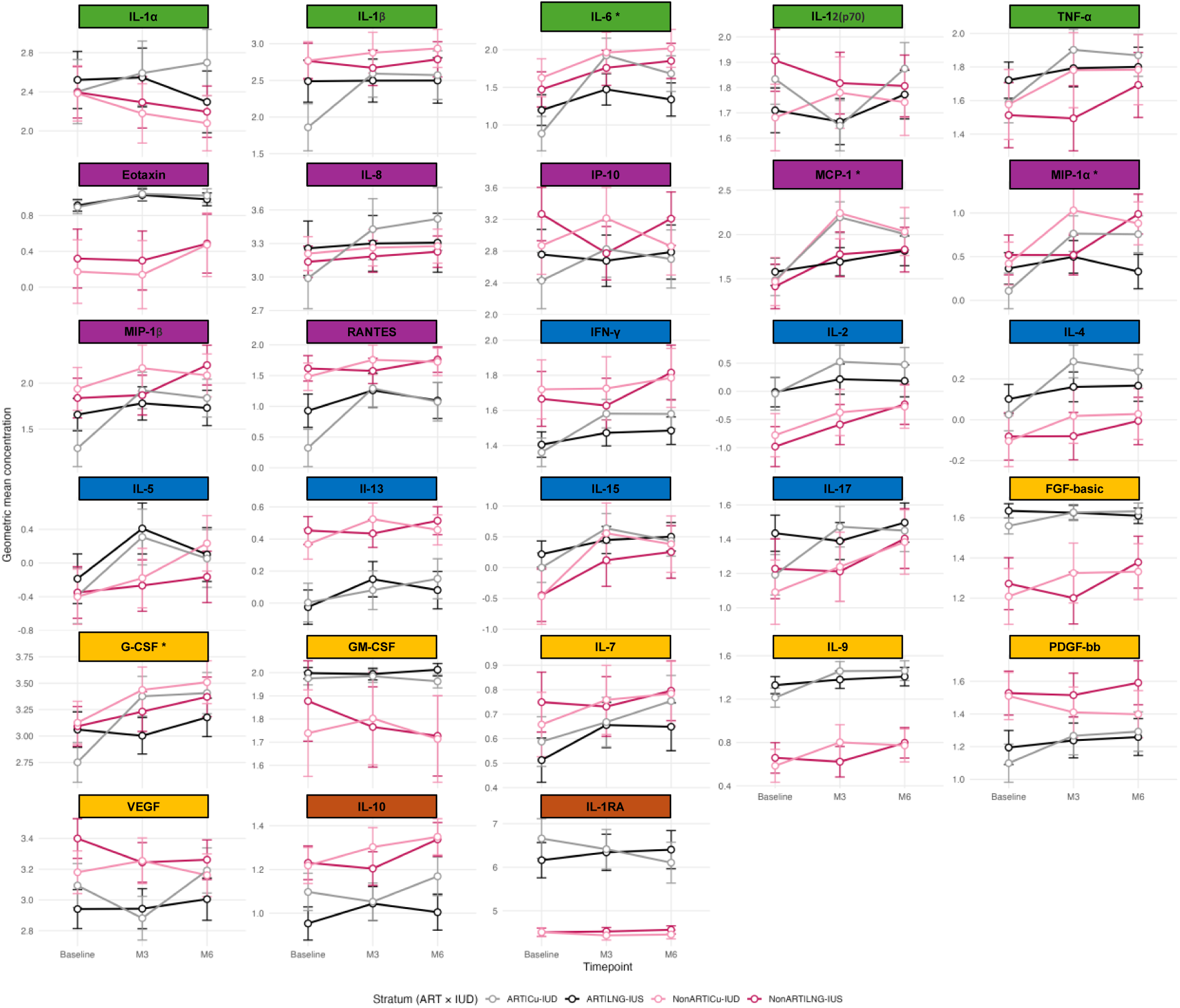
Longitudinal log_10_ geometric mean cytokine concentrations stratified by ART and IUC groups. Geometric mean concentrations of cervicovaginal cytokines and chemokines measured from baseline to month 3 (M3) to month 6 (M6) are shown on a log_10_LJscale. Each panel represents an individual cytokine, including inflammatory (green), growth factors (yellow), chemokines (purple), anti-inflammatory cytokines (red) and adaptive (blue) cytokines. Lines represent geometric mean concentrations, and error bars represent geometric standard errors. Longitudinal cytokine concentrations are shown by antiretroviral therapy (ART) stratum, and by intrauterine contraceptive (IUC) use, including ART/C-IUC (grey) versus ART/LNG-IUS (black) users, and Non-ART/C-IUC (light pink) versus Non-ART/LNG-IUS (dark pink) users. * Indicates that there was a significant interaction between time x C-IUC within the ART stratum.

Among non-ART users, few cytokines demonstrated FDR-significant overall time effects, with G-CSF, IL-10, IL-15, IL-2, IL-6, MCP-1 and MIP-1α passing FDR correction; IL-17 and IL-5 showed nominal significance prior to adjustment. Two cytokines (IP-10, p = 0.04 and MIP-1α, p = 0.02) showed nominal interaction p-values for the timeL×LCu-IUC interaction, but these did not remain significant after FDR correction. Overall, these findings indicate that IUC-dependent longitudinal cytokine patterns were not evident among non-ART users (Figure 2; Supplementary Table 5).

In contrast, among ART users, a broad set of cytokines, including Eotaxin, G-CSF, IFNLγ, ILL12(p70), IL-15, IL-17, ILL1β, IL-2, IL-4, IL-6, IL-7, IL-9, MCP-1, MIPL1α, MIPL1β, RANTES, TNF-a and VEGF, exhibited FDR-significant overall time effects, indicating consistent within-participant changes from baseline to 3 and 6 months. Four cytokines also showed IUC-dependent longitudinal patterns. FDR-significant timeL×LCu-IUC interactions were present for GLCSF, ILL6, MCPL1 and MIPL1α, with Cu-IUC users exhibiting larger increases from baseline at follow-up visits compared with the reference group (LNG-IUS). Additionally, FGF basic, ILL17, ILL1β, IL-4, and MIPL1β showed timeL×LCu-IUC interaction significance prior to adjustment, again with Cu-IUC users exhibiting larger longitudinal increases (Figure 2; Supplementary Table 5).

Together, these analyses indicate that IUC-dependent longitudinal cytokine patterns were restricted to ART users, suggesting that ART status may modify cervicovaginal immune responses to IUC insertion. Among ART users, Cu-IUC use was associated with larger increases in concentrations of cytokines across classes over time compared with LNG-IUS.

### Cytokine clustering and discrimination by IUC type

In agreement with previous analyses, ART status most strongly differentiated genital cytokine concentrations grouping using unsupervised hierarchical clustering, with ART users tending to have higher overall concentrations than non-ART women, with cytokines consistently higher despite IUC insertion (Figure 3a). As IUC insertion broadly affected cytokine levels only in ART users (Figures 1, 2), we next sought to identify the key contributors distinguishing CLIUC from LNGLIUS users within the ARTLusing subgroup. Although PERMANOVA showed no statistically significant difference in overall cytokine profile changes from baseline to 6 months between IUC groups (pL=L0.29; FigureL3b), variable importance scores (VIP) from the PLSLDA model highlighted a subset of cytokines contributing to the separation observed among ART users by IUC group when considering the change in cytokines from baseline to 6 months. ILL6 showed the strongest discriminatory influence (VIPL=L2.39), followed by VEGF (VIPL=L2.08), suggesting differential modulation of inflammatory and angiogenic pathways between CLIUC versus LNGLIUS users. Several additional cytokines with VIP values exceeding one, including ILL4 (VIPL=L1.55), MIPL1α (VIPL=L1.51), ILL1RA (VIPL=L1.31), ILL12p70 (VIPL=L1.26), ILL9 (VIPL=L1.24), MIPL1β (VIPL=L1.18), MCPL1 (VIPL=L1.07), and ILL1β (VIPL=L1.01), also contributed to group separation, indicating potential involvement of Th2Lassociated signaling and chemokineLdriven immune cell recruitment.

**Figure 3.**
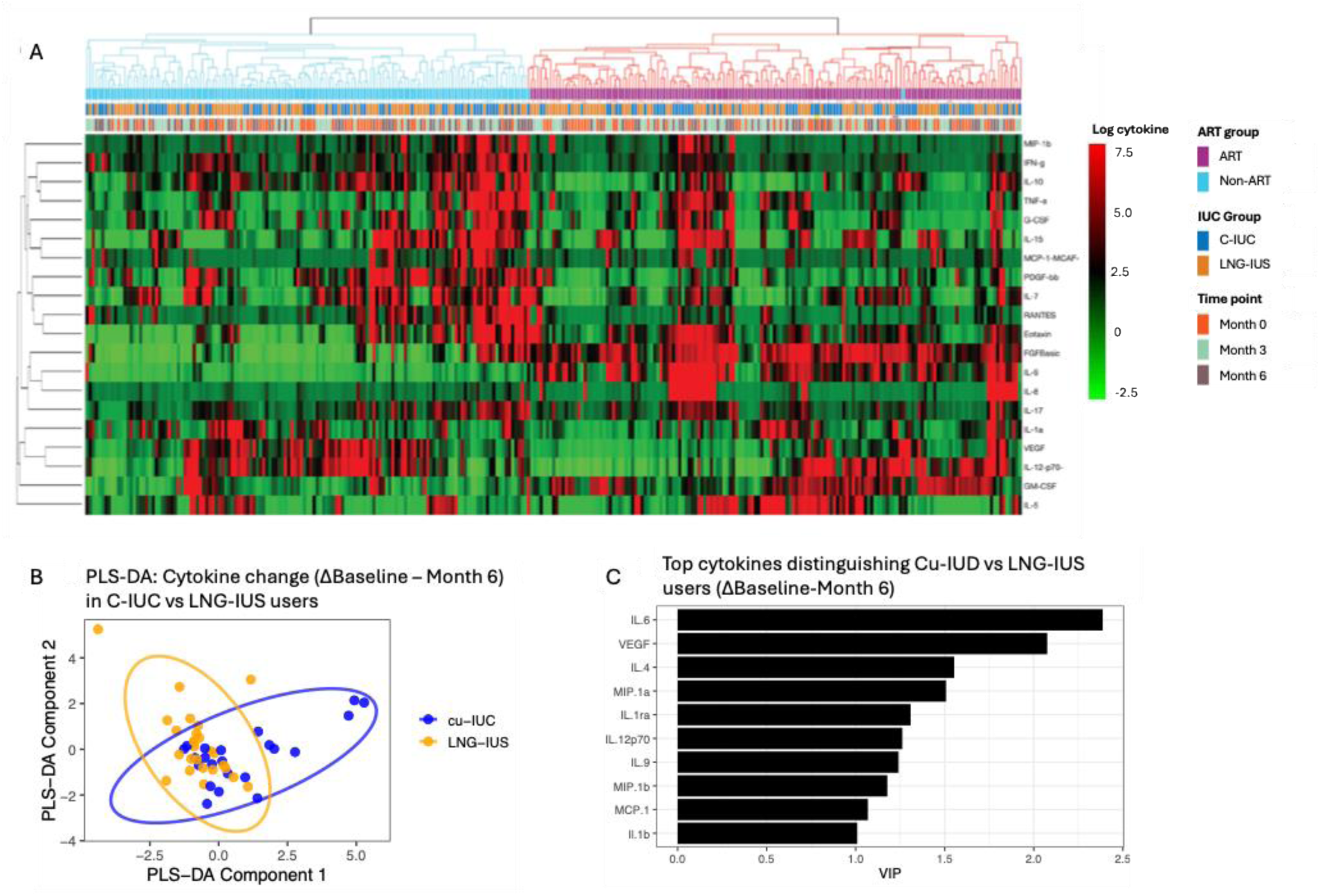
Cytokine clustering and discrimination by intrauterine contraceptive type. (A) Unsupervised hierarchical clustering of log-transformed cervicovaginal cytokine concentrations annotated by ART use [top annotation bar, ART users = purple, nonLJART users= light blue], IUC type [middle annotation bar, CLJIUC = dark blue, LNGLJIUS = orange] and time point [bottom annotation bar, baseline = orange, month 3 = green, month 6 = grey]. (B) PLSLJDA assessing changes in cytokine concentrations from baseline to monthLJ6 among ARTLJtreated participants, comparing CLJIUC and LNGLJIUS users. (C) VariableLJimportanceLJinLJprojection (VIP) scores from the PLSLJDA model, highlighting cytokines that contribute most strongly to distinguishing the two contraceptive groups based on changes in cytokine levels from baseline to monthLJ6.

## Discussion

IUCs are considered a safe and highly effective contraceptive option for WLHIV, with no evidence that IUC use accelerates HIV disease progression or increases the risk of HIV transmission to sexual partners [22,27–29]. At the same time, substantial evidence indicates that IUC insertion induces localized morphological, biochemical and inflammatory responses within the female genital tract [30–36]. Because genital inflammation is a key determinant of HIV detection in genital secretions, even among WLHIV receiving ART [19,37], we investigated whether intrauterine IUC type (C-IUC vs. LNG-IUS) differentially influences cervicovaginal cytokine responses over six months following IUC insertion in WLHIV. Based on evidence from WWOH [17,20,21], we hypothesized that Cu-IUC insertion would be associated with greater increases in pro-inflammatory cervicovaginal cytokines compared with LNG-IUS, regardless of ART status. This secondary analysis of a randomized study yielded three principal findings that clarify how ART status and IUC type appear to jointly shape genital immune responses in this population.

First, we observed marked differences in baseline genital immune profiles between ART users and nonLusers, despite similar pre-enrollment prevalences of RTIs and BV, with ART use emerging as the strongest discriminator of cytokine clusters. Untreated HIV infection was associated with substantially elevated concentrations of proLinflammatory cytokines, including ILL1α, ILL1β, ILL6, TNFLα, and chemokines, such as MIPL1α and MCPL1. This heightened mucosal inflammation is consistent with the established immunopathogenesis of untreated HIV and has been associated with vaginal epithelial disruption and HIV target cell recruitment [19,38,39]. In contrast, ART users had substantially lower genital levels of most cytokines at baseline, supporting the systematic immunomodulatory effects of viral suppression [40]. This finding is notable, as prior studies suggest that suppressive ART does not uniformly normalize cervicovaginal immune parameters [12,41,42]. Together, these findings raise important questions about how ART influences local mucosal immune homeostasis and highlight the need for longitudinal studies integrating virologic, immunologic, and clinical endpoints in the genital tract.

Second, contrary to our hypothesis, insertion of either class of IUC did not meaningfully alter genital cytokine profiles among ART nonLusers at months 3 and 6 relative to baseline, with comparable longitudinal cytokine trajectories being noted over six months between IUC types. While this contrasts with studies in WWOH, where CLIUC insertion is frequently associated with rapid increases in mucosal cytokines [16,20,43], our findings align with evidence that high baseline inflammation may attenuated subsequent mucosal immune responses to IUC exposure [21]. In women with untreated HIV, who already exhibit markedly elevated genital cytokine levels, any additional IUCLrelated inflammatory effects may be minimal relative to underlying HIVLdriven immune activation. These results suggest that baseline mucosal inflammatory state critically modifies the genital tract’s responsiveness to IUC introduction, with high pre-existing inflammation in non-ART users potentially obscuring detection of additional IUC-associated immune activation.

Third, among women using ART, C-IUC insertion was associated with broad increases in cytokines across functional classes at months 3 and 6 post-insertion relative to baseline. This finding is supported by prior randomized trials in WWOH. In sub-studies of the ECHO Trial, CLIUC use led to broad increases in cervicovaginal cytokines, while LNG-implants and DMPALIM showed little or no effect [17,20,21]. Although direct comparative data between LNG-IUS and C-IUC effects on the cervicovaginal environment remain limited, pilot observational studies similarly reported increased inflammation and microbiota perturbations with C-IUC use, and comparatively muted effects with LNGLIUS [16,44,45]. Future trials specifically designed to compare C-IUC and LNGLIUS effects on the cervicovaginal immune environment will be important to further establish causal relationships between CLIUC use and genital inflammation. With LNG-IUS use among ART users, few cytokines showed significant changes at individual follow-up time points relative to baseline, while joint tests indicated evidence of overall temporal variation, suggesting some cytokine modulation over time. Further, IL-6, IL-1β, G-CSF, FGF basic, MIP-1α, and MIP-1β showed the greatest divergence between device types in the longitudinal analyses, with steeper increases among C-IUC users. In alignment, IL-6 had the strongest influence to discriminate Cu-IUC vs. LNG-IUS users when considering the change in cytokines from baseline to month 6 post-insertion. These cytokines are centrally involved in epithelial disruption, tissue repair, leukocyte recruitment, and CCR5-mediated immune cell trafficking [17,19,39,46–48]. Collectively, cytokine patterns across month 3 versus baseline, month 6 versus baseline, and longitudinal analyses support a copper-associated amplification of genital inflammation in ART users, whereas LNGLIUS use appears to maintain a comparatively stable genital immune environment.

This study has several limitations. Although cervicovaginal cytokines are established markers of mucosal immune activation, we did not directly assess HIV transmission risk or clinical outcomes; findings should therefore be interpreted as biological signals rather than clinical endpoints. ART non-users entered the study with elevated baseline inflammation, which may have reduced sensitivity to detect additional contraceptive-associated effects. Further, post-insertion sampling beginning at 3 months may have missed transient inflammatory responses immediately following insertion. Although ART uptake has increased under Treat All guidelines, gaps in initiation and adherence persist; thus, inclusion of ART non-users remains relevant for capturing real-world variability in genital immune responses. We also note that ART regimens used in the parent trial differ from many contemporary first-line regimens. In addition, cytokines were analyzed individually without accounting for correlations between markers, potentially limiting insight into coordinated inflammatory responses. We also observed incomplete concordance across analytical approaches (visit-specific contrasts, longitudinal models, and discriminatory analyses), likely reflecting differences in the biological signals captured by each method. We also observed incomplete concordance across analytical approaches (visit-specific contrasts, longitudinal models, and discriminatory analyses), likely reflecting differences in the biological signals captured by each method. Finally, we did not evaluate whether factors such as STIs, microbiota composition, or immune cell populations mediate the relationship between IUC use and cytokine responses.

In summary, this secondary analysis leveraging samples from a longitudinal randomized study demonstrates that ART status fundamentally shapes genital mucosal immune responses to IUC in WLHIV. Among ART non-users with high baseline inflammation, neither C-IUC nor LNG-IUS insertion induced additional substantial cytokine changes. In contrast, among ART virally suppressed women with lower baseline genital inflammation, CLIUC insertion triggered distinct proLinflammatory cytokine shifts, whereas LNGLIUS use resulted in minimal immunologic change across six months. The findings among ART users are consistent with broader evidence linking copperLbased IUCs to mucosal inflammation and epithelial disruption, while LNGLreleasing systems appear comparatively immunologically quiescent [16,44,45]. While both IUCs remain safe and effective contraceptive options, our data suggest that LNGLIUS may better maintain a low-inflammatory genital environment in ART-using women, highlighting the value of integrating mucosal immunology into contraceptive research in this population.

## Acknowledgements

We thank the staff of the Gugulethu Green Clinic for their efforts and our patients for their time and trust. We also thank Dr Nicos Karasavva for his guidance on MC sample processing techniques.

## Data availability

The de-identified clinical database and variable specification files are available on DataVerse at the following link: https://dataverse.harvard.edu/dataset.xhtml?persistentId=doi:10.7910/DVN/NTN7KY. Cytokine data underlying this article will be shared on reasonable request to the corresponding author.

## Author Contributions

JAP supervised the cytokine measurements for the study, analysed the data, and wrote the manuscript; AUH analysed the data, and wrote the manuscript; MS analyzed the data and contributed to the manuscript; SZJ, HG and NH performed experiments and analysed the data; NL oversaw the running of the study and sample collection; HJ and DH assisted with preparation of the manuscript; LM and CT conceptualized and designed the study, analyzed the data, and contributed to the manuscript.

## ALT Figure Text

FIGURE 1 ALT TEXT: Multi-panel forest plots comparing changes in cervicovaginal cytokine concentrations from baseline to months 3 and 6 among women using intrauterine contraception, stratified by ART status. Panels are grouped into non-ART users and ART users, with separate plots for month 3 and month 6 comparisons. Colored point estimates with horizontal confidence intervals represent fold changes in inflammatory, chemotactic, regulatory, growth factor, and adaptive immune cytokines relative to baseline. Most cytokines cluster around no major change, although several inflammatory markers show increases over time, particularly among ART users. Vertical reference lines indicate no change from baseline.

FIGURE 2 ALT TEXT: Multi-panel line graphs showing longitudinal changes in cervicovaginal cytokine concentrations from baseline to months 3 and 6 among women using intrauterine contraception. Each panel represents a different cytokine, including inflammatory, chemotactic, adaptive immune, growth factor, and regulatory markers. Lines and symbols compare study groups across time points, with error bars indicating variability around the mean. Several inflammatory cytokines, including IL-1β, IL-6, TNF-α, MCP-1, and MIP-1α, show increases over time in some groups, while other cytokines remain relatively stable. Growth factors such as G-CSF and VEGF also demonstrate temporal variation. Overall, the figure illustrates distinct longitudinal cytokine response patterns across immune mediators and participant groups.

FIGURE 3 ALT TEXT: Multi-panel figure comparing longitudinal cytokine changes between copper intrauterine contraceptive (Cu-IUC) and levonorgestrel intrauterine system (LNG-IUS) users. Panel A shows a clustered heatmap of cytokine changes between baseline and month 6, with participants grouped by ART status, IUC type, and time point. Rows represent cytokines and columns represent individual participants. Red indicates higher cytokine expression and green indicates lower expression relative to baseline. Hierarchical clustering identifies distinct inflammatory response patterns across participants and contraceptive groups. Panel B shows a partial least squares discriminant analysis (PLS-DA) plot comparing cytokine profiles between Cu-IUC and LNG-IUS users. Individual participants cluster separately by contraceptive group, with overlapping but distinguishable distributions. Panel C displays the top cytokines contributing to differences between Cu-IUC and LNG-IUS users based on variable importance in projection (VIP) scores. IL-6 shows the highest discriminatory contribution, followed by VEGF, IL-4, MIP-1α, IL-1ra, IL-12p70, IL-9, MIP-1β, MCP-1, and IL-1β.

