## Supplemental Figures and Tables for "Genital Inflammatory Responses in Women Living with HIV Randomized to Copper or Levonorgestrel Intrauterine Contraceptives: A secondary analysis of a randomized trial": 2IUD_cytokines_supplemental_20May2026.docx

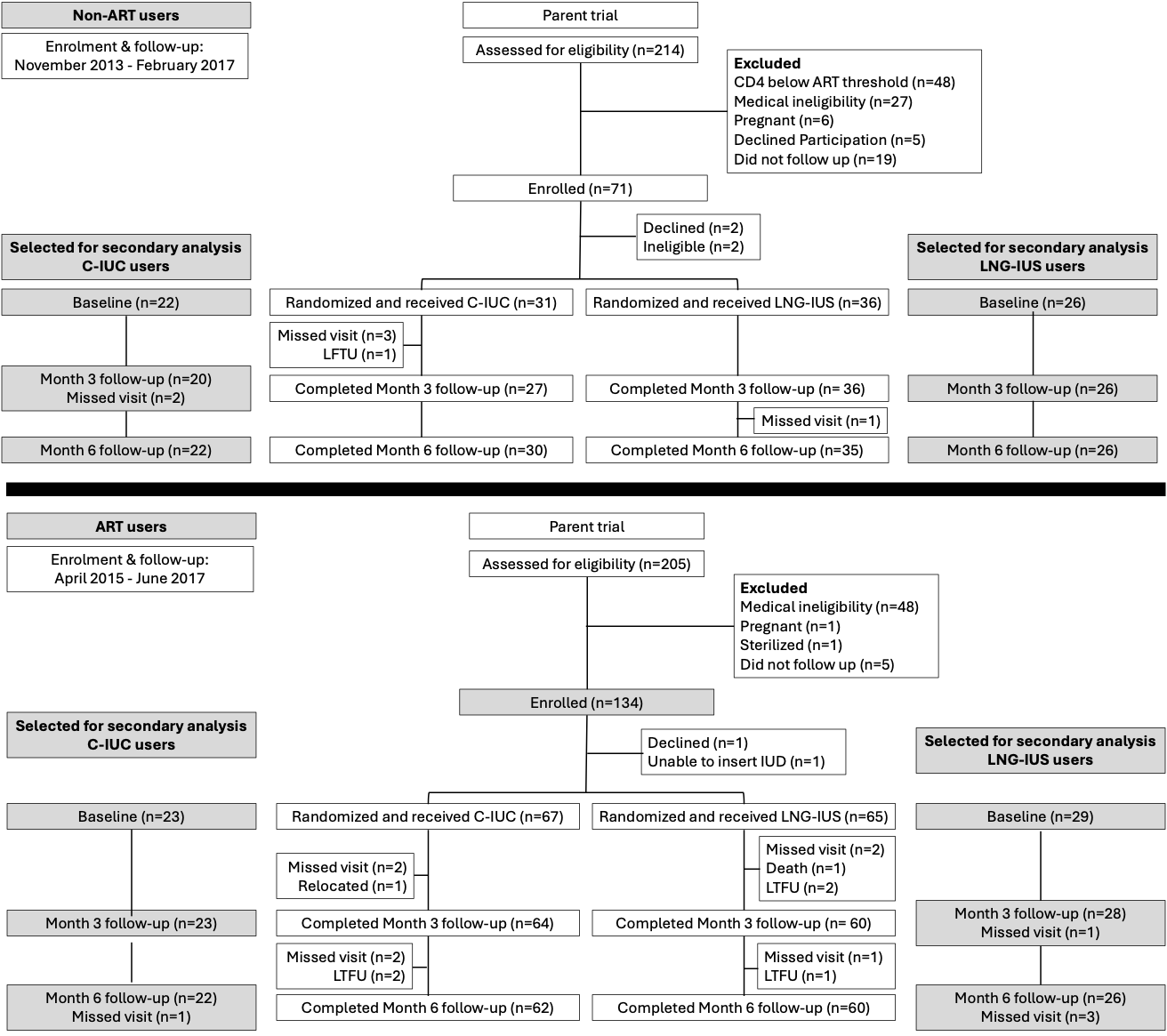

**Supplementary Figure 1. Overview of participant numbers and follow-up.** Participant numbers for the parent randomized-controlled trial are shown in white text boxes, while numbers for the secondary analysis present here are shown in the grey text boxes. C-IUC = Copper- Intrauterine contraceptive, LNG-IUS = levonorgestrel intrauterine system, LTUF: lost-to-follow-up.

**Supplementary Table 1.** Baseline genital tract cytokines by gVL detection.

| **Class** | **Cytokine**  **[median, IQR]** | **gVL > LoD (n=34)** | **gVL < LoD (n=66)** | **Adjusted p-value^#^** |
| --- | --- | --- | --- | --- |
| **Inflammatory** | IL-1α | 557.98 [326.74, 1317.28] | 583.54 [182.42, 1204.03] | 0.393 |
|  | IL-1β | 436.98 [166.69, 1396.23] | 190.01 [48.60, 941.31] | 0.083 |
|  | IL-6 | 32.44 [10.30, 113.20] | 13.94 [5.14, 38.68] | 0.052 |
|  | IL-12(p70) | 77.42 [60.00, 97.72] | 67.11 [41.42, 93.66] | 0.199 |
|  | TNF-α | 39.44 [23.82, 64.96] | 37.64 [25.43, 63.25] | 0.999 |
| **Chemokines** | Eotaxin | 6.18 [2.48, 9.71] | 7.62 [5.27, 10.09] | 0.143 |
|  | IL-8 | 1664.18 [1157.26, 3017.31] | 838.10 [356.55, 2537.44] | **0.045** |
|  | IP-10 | 2350.28 [316.10, 7855.10] | 397.67 [133.31, 1679.35] | **0.034** |
|  | MCP-1 | 28.09 [15.57, 46.34] | 35.30 [18.02, 57.65] | 0.551 |
|  | MIP-1α | 2.97 [1.90, 4.62] | 1.37 [0.90, 3.88] | **0.032** |
|  | MIP-1β | 70.03 [33.48, 172.34] | 39.34 [14.19, 90.80] | **0.047** |
|  | RANTES | 35.26 [14.07, 123.40] | 7.86 [2.24, 28.67] | **0.005** |
| **Growth factors** | FGF basic | 20.27 [15.08, 33.66] | 34.79 [26.57, 41.62] | **0.005** |
|  | G-CSF | 1964.00 [511.40-4951.00] | 804.30 [334.00-2876.00] | 0.060 |
|  | GM-CSF | 67.06 [49.65, 89.95] | 86.37 [66.41, 130.50] | **0.014** |
|  | IL-7 | 5.12 [3.72, 8.72] | 3.98 [2.43, 5.81] | **0.018** |
|  | IL-9 | 5.26 [3.34, 8.87] | 14.40 [7.47, 22.56] | **0.005** |
|  | PDGF-bb | 30.83 [16.22, 41.80] | 15.60 [11.51, 23.85] | **0.008** |
|  | VEGF | 2423.33 [1043.92, 3854.42] | 1225.48 [675.21, 2176.79] | **0.018** |
| **Adaptive** | IFN-γ | 33.63 [25.30, 63.42] | 25.07 [18.23, 41.60] | **0.032** |
|  | IL-2 | 0.02 [0.02, 2.88] | 2.34 [0.03, 3.53] | **0.018** |
|  | IL-4 | 0.91 [0.62, 1.40] | 1.04 [0.74, 1.42] | 0.393 |
|  | IL-5 | 0.50 [0.06, 3.31] | 1.01 [0.04, 3.07] | 0.963 |
|  | IL-13 | 2.50 [1.60, 3.50] | 1.18 [0.79, 2.03] | **0.005** |
|  | IL-15 | 1.20 [0.01, 6.11] | 3.43 [0.14, 9.31] | **0.049** |
|  | IL-17 | 23.07 [15.03, 34.08] | 19.65 [12.91, 34.18] | 0.490 |
| **Anti-inflammatory** | IL-10 | 16.06 [13.58, 22.30] | 11.90 [8.92, 16.32] | **0.005** |
|  | IL-1RA | 6.5x10^4^ [1.0, 10.8x10^4^] | 1088.9x10^4^ [6.5, 1088.9x10^4^] | **0.005** |

^#^P-values were adjusted for multiple comparisons using Bonferroni across all gVL detectable vs. non detectable comparisons. gVL = genital viral load, LoD = limit of detection.

**Supplementary Table 2. Baseline cohort characteristics of non-ART users randomized to C-IUC versus LNG-IUS**

|  | C-IUC | LNG-IUS | P-value |
| --- | --- | --- | --- |
| N | 22 | 26 |  |
| Age, years (median; IQR) | 31 [28, 33] | 30 [26, 34] | 0.973 |
| In relationship (n/N; %) | 19 (95.0) | 25 (96.2) | 0.920 |
| Reproductive tract infections screening (n; %)^@^  *C. trachomatis*  *N. gonnorhoea*  *T. vaginalis*  HSV-2 [shedding]  HSV-2 [IgG]  Bacterial vaginosis* | 1 (4.5)  1 (4.5)  2 (9.1)  2 (9.1)  18 (81.8)  2 (9.1) | 2 (7.7)  1 (2.7)  1 (2.7)  2 (7.7)  23 (88.5)  6 (23.1) | 1.000  1.000  0.881  0.952  0.445  0.364 |
| Cytokines^#^ |  |  |  |
| IL-1α | 893.18 [507.82, 1358.39] | 716.36 [266.18, 1363.92] | 0.944 |
| IL-1β | 926.59 [189.21, 1680.58] | 800.05 [212.37, 1357.17] | 0.820 |
| IL-6 | 59.99 [15.01, 139.81] | 42.80 [12.49, 113.20] | 0.593 |
| IL-12(p70) | 73.72 [58.36, 85.18] | 80.58 [64.58, 105.29] | 0.185 |
| TNF-α | 33.92 [14.86, 86.65] | 42.17 [12.05, 62.62] | 0.932 |
| Eotaxin | 2.70 [0.36, 10.77] | 6.41 [2.22, 9.52] | 0.393 |
| IL-8 | 1776.14 [1220.22, 3044.70] | 1372.56 [721.44, 2926.60] | 0.522 |
| IP-10 | 773.75 [258.08, 3500.28] | 3603.36 [461.83, 10328.39] | 0.331 |
| MCP-1 | 20.39 [15.29, 69.75] | 23.42 [12.48, 52.24] | 0.822 |
| MIP-1α | 3.06 [1.93, 4.62] | 3.30 [1.95, 5.50] | 0.422 |
| MIP-1β | 93.06 [44.74, 198.59] | 70.03 [43.85, 227.65] | 0.788 |
| RANTES | 31.01 [13.83, 68.15] | 39.33 [14.40, 111.84] | 0.432 |
| FGF basic | 18.26 [14.46, 24.41] | 21.00 [14.14, 25.73] | 0.710 |
| G-CSF | 1892.87 [831.24, 5499.78] | 1999.74 [435.51, 4602.33] | 0.710 |
| GM-CSF | 59.81 [46.40, 75.96] | 69.59 [56.63, 83.72] | 0.185 |
| IL-7 | 5.22 [3.73, 6.80] | 4.70 [3.66, 9.64] | 0.812 |
| IL-9 | 4.65 [3.52, 6.56] | 5.26 [3.30, 6.62] | 0.788 |
| PDGF-bb | 31.04 [23.55, 40.02] | 26.68 [16.52, 51.05] | 0.692 |
| VEGF | 2084.47 [1156.56, 3127.18] | 2423.33 [1467.57, 4170.32] | 0.379 |
| IFN-γ | 46.57 [25.95, 64.14] | 49.83 [31.45, 77.96] | 0.661 |
| IL-2 | 0.04 [0.01, 1.70] | 0.01 [0.01, 2.81] | 0.600 |
| IL-4 | 0.78 [0.60, 1.14] | 0.88 [0.49, 1.38] | 0.885 |
| IL-5 | 0.84 [0.13, 2.23] | 0.50 [0.10, 5.24] | 0.860 |
| IL-13 | 2.50 [1.91, 3.46] | 2.83 [2.02, 3.57] | 0.521 |
| IL-15 | 1.59 [0.01, 7.59] | 3.30 [0.01, 4.72] | 0.897 |
| IL-17 | 20.27 [13.81, 34.11] | 29.62 [14.72, 42.55] | 0.385 |
| IL-10 | 16.42 [14.22, 20.16] | 16.31 [14.48, 20.90] | 0.804 |
| IL-1RA | 46123 [9586, 105827] | 62229 [9416, 95335] | 0.664 |

^@^Women were treated for RTIs prior to enrollment. ^*^Determined by OSOM® BV Blue ^#^P-values were adjusted for multiple comparisons using Bonferroni across all Cu-IUC vs. LNG-IUS comparisons.

**Supplementary Table 3. Baseline cohort characteristics of ART users randomized to C-IUC versus LNG-IUS**

|  | C-IUC | LNG-IUS | P-value |
| --- | --- | --- | --- |
| N | 23 | 29 |  |
| Age, years (median; IQR) | 30 [27, 33] | 31 [28, 34] | 0.301 |
| In relationship (n/N; %) | 21 (91.3) | 27 (93.1) | 0.820 |
| Reproductive tract infections screening (n; %)^@^  *C. trachomatis*  *N. gonnorhoea*  *T. vaginalis*  HSV-2 [shedding]  HSV-2 [IgG]^a^  Bacterial vaginosis* | 1 (4.3)  2 (8.7)  1 (4.3)  1 (4.3)  20 (87.0)  2 (8.9) | 2 (6.9)  2 (6.9)  5 (17.2)  1 (3.4)  27 (93.1)  5 (17.2) | 1.000  1.000  0.313  1.000  0.304  0.626 |
| Cytokines^#^ |  |  |  |
| IL-1α | 389.00 [218.53, 879.59] | 523.38 [201.52, 1054.44] | 0.724 |
| IL-1β | 67.60 [24.12, 264.05] | 245.68 [49.54, 829.64] | 0.145 |
| IL-6 | 6.14 [4.01, 14.30] | 14.09 [6.73, 22.92] | 0.168 |
| IL-12(p70) | 90.36 [56.26, 107.94] | 53.10 [38.66, 78.76] | 0.168 |
| TNF-α | 36.94 [25.75, 47.89] | 51.26 [29.78, 72.86] | 0.200 |
| Eotaxin | 7.26 [5.80, 9.74] | 8.19 [6.91, 9.90] | 0.643 |
| IL-8 | 584.70 [261.76, 1405.64] | 1194.96 [687.31, 2346.99] | 0.168 |
| IP-10 | 275.44 [71.01, 1045.08] | 435.10 [102.29, 1462.02] | 0.363 |
| MCP-1 | 35.57 [17.80, 42.42] | 38.15 [21.02, 73.41] | 0.363 |
| MIP-1α | 0.96 [0.69, 1.49] | 1.50 [1.12, 3.14] | 0.145 |
| MIP-1β | 16.30 [8.62, 39.07] | 39.17 [17.88, 71.91] | 0.145 |
| RANTES | 2.24 [0.84, 7.35] | 8.21 [3.00, 32.53] | 0.126 |
| FGF basic | 36.18 [31.70, 39.99] | 42.22 [35.41, 45.72] | 0.145 |
| G-CSF | 601.06 [241.74, 1701.23] | 982.49 [541.03, 3014.88] | 0.200 |
| GM-CSF | 107.07 [74.29, 124.04] | 100.79 [73.60, 131.25] | 0.661 |
| IL-7 | 4.28 [2.59, 6.34] | 3.49 [2.17, 4.84] | 0.305 |
| IL-9 | 15.86 [13.00, 21.10] | 19.54 [16.37, 31.83] | 0.182 |
| PDGF-bb | 13.70 [10.16, 19.10] | 14.91 [10.17, 24.91] | 0.402 |
| VEGF | 1459.85 [1042.90, 2033.72] | 867.84 [512.32, 1522.46] | 0.168 |
| IFN-γ | 22.50 [17.04, 26.85] | 25.62 [19.38, 33.77] | 0.352 |
| IL-2 | 1.92 [0.35, 3.53] | 2.41 [0.38, 3.83] | 0.783 |
| IL-4 | 1.04 [0.81, 1.26] | 1.27 [0.94, 1.73] | 0.207 |
| IL-5 | 0.39 [0.04, 2.92] | 1.04 [0.04, 3.18] | 0.713 |
| IL-13 | 1.16 [0.74, 1.78] | 0.93 [0.71, 1.24] | 0.366 |
| IL-15 | 1.45 [0.14, 4.19] | 2.38 [0.14, 9.42] | 0.436 |
| IL-17 | 14.59 [12.07, 20.35] | 22.80 [17.31, 31.88] | 0.056 |
| IL-10 | 13.65 [8.41, 19.49] | 10.06 [6.72, 12.03] | 0.363 |
| IL-1RA | 10889000 [10889000, 10889000] | 10889000 [67721, 10889000] | 0.207 |

^@^Women were treated for RTIs prior to enrollment. ^*^Determined by OSOM® BV Blue ^#^P-values were adjusted for multiple comparisons using Bonferroni across all Cu-IUC vs. LNG-IUS comparisons. ^a^ missing data n=3

**Supplementary Table 4.** Log_10_ fold-changes (Geometric Mean Ratio, GMR) in cytokines from month 3 [M3] compared to baseline, or from month 6 [M6] compared to baseline, by IUC group for Non-ART and ART users.

|  |  |  | **Non-ART users** | | **ART users** | |
| --- | --- | --- | --- | --- | --- | --- |
| **Cytokine** | **Contrast** | **IUC group** | **Log_10_ GMR (95% CI)** | **p-value (adjusted)** | **Log_10_ GMR (95% CI)** | **p-value (adjusted)** |
| Eotaxin | M3 / Baseline | C-IUC | -0.03 (-0.55, 0.48) | 0.895 | 0.15 (0.04, 0.25) | **0.007 (0.026)** |
| Eotaxin | M6 / Baseline | C-IUC | 0.30 (-0.20, 0.80) | 0.238 | 0.13 (0.02, 0.23) | **0.019** (0.060) |
| IL-1α | M3 / Baseline | LNG-IUS | -0.02 (-0.49, 0.44) | 0.925 | 0.12 (0.02, 0.21) | **0.017**(0.056) |
| IL-1β | M6 / Baseline | LNG-IUS | 0.17 (-0.30, 0.63) | 0.477 | 0.07 (-0.03, 0.16) | 0.169 |
| IL-6 | M3 / Baseline | C-IUC | 0.12 (-0.09, 0.32) | 0.259 | 0.07 (0.01, 0.12) | **0.021** (0.063) |
| IL-12(p70) | M6 / Baseline | C-IUC | 0.12 (-0.07, 0.32) | 0.215 | 0.07 (0.02, 0.13) | **0.014 (0.048)** |
| TNF-α | M3 / Baseline | LNG-IUS | -0.07 (-0.25, 0.11) | 0.438 | -0.01 (-0.06, 0.04) | 0.725 |
| Eotaxin | M6 / Baseline | LNG-IUS | 0.11 (-0.08, 0.29) | 0.249 | -0.02 (-0.08, 0.03) | 0.358 |
| IL-8 | M3 / Baseline | C-IUC | 0.31 (0.01, 0.61) | **0.040** (0.225) | 0.63 (0.36, 0.89) | **<0.0001 (0.0002)** |
| IP-10 | M6 / Baseline | C-IUC | 0.38 (0.10, 0.67) | **0.009** (0.125) | 0.66 (0.38, 0.93) | **<0.0001 (0.0002)** |
| MCP-1 | M3 / Baseline | LNG-IUS | 0.14 (-0.13, 0.41) | 0.300 | -0.06 (-0.30, 0.18) | 0.633 |
| MIP-1α | M6 / Baseline | LNG-IUS | 0.28 (0.01, 0.54) | **0.040** (0.225) | 0.12 (-0.13, 0.36) | 0.356 |
| MIP-1β | M3 / Baseline | C-IUC | 0.06 (-0.21, 0.34) | 0.646 | 0.01 (-0.03, 0.05) | 0.629 |
| RANTES | M6 / Baseline | C-IUC | -0.02 (-0.29, 0.24) | 0.852 | -0.01 (-0.05, 0.03) | 0.526 |
| FGF basic | M3 / Baseline | LNG-IUS | -0.11 (-0.36, 0.13) | 0.367 | -0.00 (-0.04, 0.03) | 0.82 |
| G-CSF | M6 / Baseline | LNG-IUS | -0.15 (-0.40, 0.10) | 0.227 | 0.01 (-0.02, 0.05) | 0.425 |
| GM-CSF | M3 / Baseline | C-IUC | 0.01 (-0.24, 0.25) | 0.966 | 0.22 (0.11, 0.34) | **0.0002 (0.002)** |
| IL-7 | M6 / Baseline | C-IUC | 0.07 (-0.17, 0.30) | 0.585 | 0.22 (0.10, 0.34) | **0.0003 (0.002)** |
| IL-9 | M3 / Baseline | LNG-IUS | -0.04 (-0.26, 0.18) | 0.732 | 0.07 (-0.04, 0.17) | 0.202 |
| PDGF-bb | M6 / Baseline | LNG-IUS | 0.15 (-0.07, 0.37) | 0.181 | 0.08 (-0.03, 0.19) | 0.140 |
| VEGF | M3 / Baseline | C-IUC | 0.08 (-0.04, 0.21) | 0.172 | -0.04 (-0.17, 0.08) | 0.463 |
| IFN-γ | M6 / Baseline | C-IUC | 0.13 (0.01, 0.25) | **0.029 (**0.199) | 0.07 (-0.05, 0.19) | 0.251 |
| IL-2 | M3 / Baseline | LNG-IUS | -0.03 (-0.14, 0.08) | 0.633 | 0.09 (-0.02, 0.20) | 0.100 |
| IL-4 | M6 / Baseline | LNG-IUS | 0.11 (-0.00, 0.22) | 0.053 | 0.05 (-0.06, 0.16) | 0.362 |
| IL-5 | M3 / Baseline | C-IUC | 0.10 (-0.09, 0.29) | 0.313 | -0.18 (-0.32, -0.04) | **0.011 (0.040)** |
| IL-13 | M6 / Baseline | C-IUC | 0.06 (-0.13, 0.25) | 0.519 | 0.04 (-0.10, 0.18) | 0.567 |
| IL-15 | M3 / Baseline | LNG-IUS | -0.09 (-0.26, 0.08) | 0.304 | -0.04 (-0.17, 0.08) | 0.490 |
| IL-17 | M6 / Baseline | LNG-IUS | -0.10 (-0.27, 0.07) | 0.247 | 0.06 (-0.07, 0.19) | 0.344 |
| IL-10 | M3 / Baseline | C-IUC | 0.16 (0.02, 0.29) | **0.027 (**0.199) | 0.08 (-0.09, 0.25) | 0.370 |
| IL-1RA | M6 / Baseline | C-IUC | 0.09 (-0.04, 0.22) | 0.188 | 0.15 (-0.02, 0.32) | 0.092 |
| IL-13 | M3 / Baseline | LNG-IUS | -0.02 (-0.14, 0.11) | 0.768 | 0.17 (0.02, 0.33) | **0.027** (0.073) |
| IL-13 | M6 / Baseline | LNG-IUS | 0.06 (-0.06, 0.18) | 0.326 | 0.11 (-0.05, 0.27) | 0.185 |
| IL-15 | M3 / Baseline | C-IUC | 1.02 (0.35, 1.69) | **0.003** (0.092) | 0.64 (0.30, 0.98) | **0.0003 (0.002)** |
| IL-15 | M6 / Baseline | C-IUC | 0.84 (0.20, 1.49) | **0.011** (0.126) | 0.43 (0.09, 0.78) | **0.015 (0.049)** |
| IL-15 | M3 / Baseline | LNG-IUS | 0.57 (-0.03, 1.17) | 0.063 | 0.23 (-0.08, 0.54) | 0.141 |
| IL-15 | M6 / Baseline | LNG-IUS | 0.70 (0.10, 1.31) | **0.022** (0.196) | 0.28 (-0.03, 0.60) | 0.078 |
| IL-17 | M3 / Baseline | C-IUC | 0.15 (-0.13, 0.42) | 0.288 | 0.28 (0.11, 0.45) | **0.001 (0.007)** |
| IL-17 | M6 / Baseline | C-IUC | 0.29 (0.03, 0.56) | **0.031** (0.199) | 0.26 (0.09, 0.43) | **0.003 (0.015)** |
| IL-17 | M3 / Baseline | LNG-IUS | -0.02 (-0.26, 0.23) | 0.899 | -0.05 (-0.20, 0.11) | 0.554 |
| IL-17 | M6 / Baseline | LNG-IUS | 0.18 (-0.07, 0.42) | 0.160 | 0.06 (-0.09, 0.22) | 0.428 |
| IL-1a | M3 / Baseline | C-IUC | -0.20 (-0.62, 0.21) | 0.329 | 0.19 (-0.27, 0.65) | 0.419 |
| IL-1a | M6 / Baseline | C-IUC | -0.30 (-0.70, 0.10) | 0.134 | 0.30 (-0.18, 0.77) | 0.216 |
| IL-1a | M3 / Baseline | LNG-IUS | -0.10 (-0.47, 0.27) | 0.584 | 0.03 (-0.39, 0.44) | 0.905 |
| IL-1a | M6 / Baseline | LNG-IUS | -0.20 (-0.57, 0.17) | 0.290 | -0.22 (-0.66, 0.21) | 0.303 |
| IL-1b | M3 / Baseline | C-IUC | 0.11 (-0.27, 0.49) | 0.568 | 0.73 (0.28, 1.19) | **0.002 (0.009)** |
| IL-1b | M6 / Baseline | C-IUC | 0.17 (-0.20, 0.53) | 0.370 | 0.71 (0.25, 1.17) | **0.003 (0.013)** |
| IL-1b | M3 / Baseline | LNG-IUS | -0.10 (-0.44, 0.25) | 0.579 | 0.01 (-0.40, 0.42) | 0.967 |
| IL-1b | M6 / Baseline | LNG-IUS | 0.02 (-0.32, 0.36) | 0.902 | 0.01 (-0.41, 0.43) | 0.963 |
| IL-1ra | M3 / Baseline | C-IUC | -0.07 (-0.22, 0.08) | 0.328 | -0.25 (-0.89, 0.40) | 0.448 |
| IL-1ra | M6 / Baseline | C-IUC | -0.05 (-0.20, 0.09) | 0.460 | -0.55 (-1.21, 0.10) | 0.095 |
| IL-1ra | M3 / Baseline | LNG-IUS | 0.02 (-0.12, 0.15) | 0.806 | 0.18 (-0.40, 0.76) | 0.542 |
| IL-1ra | M6 / Baseline | LNG-IUS | 0.06 (-0.08, 0.19) | 0.412 | 0.24 (-0.35, 0.84) | 0.423 |
| IL-2 | M3 / Baseline | C-IUC | 0.41 (-0.16, 0.97) | 0.155 | 0.57 (0.15, 0.98) | **0.008 (0.031)** |
| IL-2 | M6 / Baseline | C-IUC | 0.51 (-0.03, 1.05) | 0.066 | 0.51 (0.09, 0.94) | **0.018** (0.056) |
| IL-2 | M3 / Baseline | LNG-IUS | 0.39 (-0.11, 0.90) | 0.125 | 0.23 (-0.15, 0.60) | 0.228 |
| IL-2 | M6 / Baseline | LNG-IUS | 0.75 (0.25, 1.25) | **0.004** (0.092) | 0.20 (-0.19, 0.59) | 0.308 |
| IL-4 | M3 / Baseline | C-IUC | 0.12 (-0.06, 0.31) | 0.187 | 0.26 (0.15, 0.37) | **<0.0001 (0.0003)** |
| IL-4 | M6 / Baseline | C-IUC | 0.13 (-0.04, 0.31) | 0.140 | 0.21 (0.10, 0.33) | **0.0004 (0.003)** |
| IL-4 | M3 / Baseline | LNG-IUS | 0.00 (-0.16, 0.17) | 0.986 | 0.06 (-0.04, 0.16) | 0.251 |
| IL-4 | M6 / Baseline | LNG-IUS | 0.08 (-0.09, 0.24) | 0.359 | 0.07 (-0.04, 0.17) | 0.219 |
| IL-5 | M3 / Baseline | C-IUC | 0.22 (-0.26, 0.70) | 0.365 | 0.70 (0.23, 1.16) | **0.004 (0.017)** |
| IL-5 | M6 / Baseline | C-IUC | 0.63 (0.17, 1.10) | **0.008** (0.125) | 0.44 (-0.03, 0.92) | 0.068 |
| IL-5 | M3 / Baseline | LNG-IUS | 0.08 (-0.35, 0.51) | 0.706 | 0.60 (0.17, 1.02) | **0.006 (0.025)** |
| IL-5 | M6 / Baseline | LNG-IUS | 0.18 (-0.25, 0.62) | 0.397 | 0.29 (-0.15, 0.72) | 0.192 |
| IL-6 | M3 / Baseline | C-IUC | 0.34 (-0.03, 0.71) | 0.074 | 1.04 (0.72, 1.36) | **<0.0001 (<0.001)** |
| IL-6 | M6 / Baseline | C-IUC | 0.39 (0.03, 0.75) | **0.032** (0.199) | 0.80 (0.47, 1.13) | **<0.001 (0.0002)** |
| IL-6 | M3 / Baseline | LNG-IUS | 0.29 (-0.04, 0.62) | 0.089 | 0.28 (-0.02, 0.57) | 0.064 |
| IL-6 | M6 / Baseline | LNG-IUS | 0.38 (0.05, 0.71) | **0.025** (0.199) | 0.14 (-0.16, 0.44) | 0.352 |
| IL-7 | M3 / Baseline | C-IUC | 0.10 (-0.09, 0.29) | 0.304 | 0.08 (-0.06, 0.22) | 0.273 |
| IL-7 | M6 / Baseline | C-IUC | 0.13 (-0.06, 0.31) | 0.182 | 0.17 (0.02, 0.31) | **0.026** (0.073) |
| IL-7 | M3 / Baseline | LNG-IUS | -0.02 (-0.19, 0.15) | 0.836 | 0.14 (0.01, 0.27) | **0.030** (0.081) |
| IL-7 | M6 / Baseline | LNG-IUS | 0.05 (-0.13, 0.22) | 0.595 | 0.14 (0.00, 0.27) | **0.045** (0.112) |
| IL-8 | M3 / Baseline | C-IUC | 0.05 (-0.17, 0.27) | 0.642 | 0.44 (0.05, 0.83) | **0.027** (0.073) |
| IL-8 | M6 / Baseline | C-IUC | 0.07 (-0.15, 0.28) | 0.537 | 0.53 (0.14, 0.92) | **0.009 (0.032)** |
| IL-8 | M3 / Baseline | LNG-IUS | 0.05 (-0.15, 0.25) | 0.634 | 0.04 (-0.31, 0.39) | 0.803 |
| IL-8 | M6 / Baseline | LNG-IUS | 0.09 (-0.11, 0.29) | 0.369 | 0.05 (-0.31, 0.41) | 0.781 |
| IL-9 | M3 / Baseline | C-IUC | 0.22 (-0.01, 0.44) | 0.056 | 0.24 (0.12, 0.37) | **0.0002 (0.002)** |
| IL-9 | M6 / Baseline | C-IUC | 0.19 (-0.03, 0.40) | 0.086 | 0.25 (0.12, 0.37) | **0.0002 (0.002)** |
| IL-9 | M3 / Baseline | LNG-IUS | -0.04 (-0.24, 0.16) | 0.72 | 0.05 (-0.06, 0.16) | 0.37 |
| IL-9 | M6 / Baseline | LNG-IUS | 0.14 (-0.06, 0.34) | 0.171 | 0.08 (-0.04, 0.19) | 0.181 |
| IP-10 | M3 / Baseline | C-IUC | 0.34 (-0.19, 0.87) | 0.203 | 0.40 (-0.10, 0.90) | 0.12 |
| IP-10 | M6 / Baseline | C-IUC | -0.01 (-0.52, 0.50) | 0.971 | 0.27 (-0.24, 0.78) | 0.292 |
| IP-10 | M3 / Baseline | LNG-IUS | -0.49 (-0.97, -0.02) | **0.042** (0.226) | -0.08 (-0.53, 0.37) | 0.73 |
| IP-10 | M6 / Baseline | LNG-IUS | -0.06 (-0.54, 0.42) | 0.806 | 0.03 (-0.44, 0.50) | 0.897 |
| MCP-1 | M3 / Baseline | C-IUC | 0.77 (0.38, 1.17) | **0.0002 (0.024)** | 0.71 (0.46, 0.96) | **<0.0001 (<0.0001)** |
| MCP-1 | M6 / Baseline | C-IUC | 0.57 (0.18, 0.95) | **0.004** (0.092) | 0.52 (0.27, 0.77) | **<0.0001 (0.001)** |
| MCP-1 | M3 / Baseline | LNG-IUS | 0.36 (0.01, 0.72) | **0.046** (0.236) | 0.11 (-0.11, 0.34) | 0.314 |
| MCP-1 | M6 / Baseline | LNG-IUS | 0.42 (0.06, 0.77) | **0.023** (0.196) | 0.24 (0.01, 0.47) | **0.043** (0.108) |
| MIP-1a | M3 / Baseline | C-IUC | 0.61 (0.25, 0.97) | **0.001** (0.064) | 0.66 (0.37, 0.94) | **<0.0001 (0.003)** |
| MIP-1a | M6 / Baseline | C-IUC | 0.46 (0.11, 0.81) | **0.010** (0.125) | 0.65 (0.36, 0.94) | **<0.0001 (0.003)** |
| MIP-1a | M3 / Baseline | LNG-IUS | -0.00 (-0.32, 0.32) | 0.996 | 0.13 (-0.13, 0.39) | 0.314 |
| MIP-1a | M6 / Baseline | LNG-IUS | 0.47 (0.15, 0.79) | **0.005** (0.092) | -0.04 (-0.30, 0.23) | 0.792 |
| MIP-1b | M3 / Baseline | C-IUC | 0.23 (-0.12, 0.57) | 0.192 | 0.63 (0.35, 0.91) | **<0.0001 (0.003)** |
| MIP-1b | M6 / Baseline | C-IUC | 0.15 (-0.18, 0.48) | 0.378 | 0.55 (0.26, 0.83) | **0.0003 (0.002)** |
| MIP-1b | M3 / Baseline | LNG-IUS | 0.03 (-0.27, 0.34) | 0.838 | 0.12 (-0.13, 0.38) | 0.340 |
| MIP-1b | M6 / Baseline | LNG-IUS | 0.36 (0.05, 0.66) | **0.022** (0.196) | 0.07 (-0.19, 0.33) | 0.579 |
| PDGF-bb | M3 / Baseline | C-IUC | -0.10 (-0.31, 0.11) | 0.352 | 0.17 (0.00, 0.33) | **0.047** (0.114) |
| PDGF-bb | M6 / Baseline | C-IUC | -0.11 (-0.32, 0.09) | 0.281 | 0.19 (0.03, 0.36) | **0.025** (0.072) |
| PDGF-bb | M3 / Baseline | LNG-IUS | -0.01 (-0.20, 0.18) | 0.898 | 0.04 (-0.11, 0.19) | 0.564 |
| PDGF-bb | M6 / Baseline | LNG-IUS | 0.06 (-0.13, 0.25) | 0.509 | 0.06 (-0.09, 0.22) | 0.409 |
| RANTES | M3 / Baseline | C-IUC | 0.27 (-0.05, 0.60) | 0.098 | 0.97 (0.54, 1.40) | **<0.0001 (0.003)** |
| RANTES | M6 / Baseline | C-IUC | 0.24 (-0.07, 0.56) | 0.129 | 0.75 (0.31, 1.19) | **0.0009 (0.005)** |
| RANTES | M3 / Baseline | LNG-IUS | -0.04 (-0.33, 0.25) | 0.784 | 0.33 (-0.06, 0.72) | 0.094 |
| RANTES | M6 / Baseline | LNG-IUS | 0.14 (-0.15, 0.44) | 0.335 | 0.17 (-0.23, 0.56) | 0.411 |
| TNF-a | M3 / Baseline | C-IUC | 0.20 (-0.10, 0.51) | 0.187 | 0.31 (0.14, 0.49) | **0.0005 (0.003)** |
| TNF-a | M6 / Baseline | C-IUC | 0.21 (-0.09, 0.50) | 0.167 | 0.28 (0.11, 0.45) | **0.002 (0.009)** |
| TNF-a | M3 / Baseline | LNG-IUS | -0.02 (-0.29, 0.26) | 0.891 | 0.07 (-0.08, 0.23) | 0.364 |
| TNF-a | M6 / Baseline | LNG-IUS | 0.18 (-0.09, 0.45) | 0.195 | 0.08 (-0.08, 0.24) | 0.328 |
| VEGF | M3 / Baseline | C-IUC | 0.07 (-0.13, 0.28) | 0.471 | -0.21 (-0.41, -0.01) | **0.039** (0.102) |
| VEGF | M6 / Baseline | C-IUC | -0.02 (-0.22, 0.18) | 0.843 | 0.10 (-0.11, 0.30) | 0.344 |
| VEGF | M3 / Baseline | LNG-IUS | -0.16 (-0.34, 0.03) | 0.094 | 0.00 (-0.18, 0.18) | 0.980 |
| VEGF | M6 / Baseline | LNG-IUS | -0.14 (-0.32, 0.04) | 0.136 | 0.06 (-0.12, 0.25) | 0.496 |

Log_10_ GMRs and 95% confidence interval (CI), raw p-values and p‑values adjusted for multiple comparisons using the Benjamini–Hochberg false discovery rate procedure, applied separately within ART strata across all cytokines and time‑point contrasts (M3 vs baseline and M6 vs baseline), are shown. P-values < 0.05 are highlighted in bold.

|  | **Non‑ART users** | | **ART users** | |
| --- | --- | --- | --- | --- |
| Cytokine | Time (adjusted) | Time × C‑IUC (adjusted) | Time (adjusted) | Time × C‑IUC (adjusted) |
| Eotaxin | 0.260 | 0.899 | **0.001 (0.002)** | 0.713 |
| FGF basic | 0.203 | 0.329 | 0.276 | **0.035** (0.109) |
| G-CSF | **0.004 (0.024)** | 0.689 | **0.0002 (0.0009)** | **0.0007 (0.018)** |
| GM-CSF | 0.611 | 0.618 | 0.978 | 0.317 |
| IFN-g | 0.267 | 0.736 | **0.0002 (0.0009)** | 0.102 |
| IL-10 | **0.011 (0.043)** | 0.375 | 0.339 | 0.131 |
| IL-12(p70) | 0.920 | 0.291 | **0.003 (0.007)** | 0.298 |
| IL-13 | 0.198 | 0.141 | **0.046** (0.065) | 0.485 |
| IL-15 | **0.0005 (0.007)** | 0.599 | **0.0006 (0.0015)** | 0.209 |
| IL-17 | **0.035** (0.122) | 0.661 | **0.019 (0.030)** | **0.018** (0.086) |
| IL-1a | 0.187 | 0.910 | 0.787 | 0.263 |
| IL-1b | 0.712 | 0.712 | **0.028 (0.043)** | **0.033** (0.109) |
| IL-1ra | 0.804 | 0.503 | 0.765 | 0.206 |
| IL-2 | **0.004 (0.023)** | 0.746 | **0.010 (0.018)** | 0.414 |
| IL-4 | 0.230 | 0.621 | **<0.0001 (0.0007)** | **0.029** (0.109) |
| IL-5 | **0.040** (0.123) | 0.361 | **0.0005 (0.0013)** | 0.889 |
| IL-6 | **0.005 (0.025)** | 0.979 | **<0.0001 (<0.0001)** | **0.001 (0.018)** |
| IL-7 | 0.406 | 0.650 | **0.008 (0.015)** | 0.633 |
| IL-8 | 0.560 | 0.980 | 0.069 | 0.160 |
| IL-9 | 0.092 | 0.213 | **0.0003 (0.0009)** | 0.051 |
| IP-10 | 0.0915 | **0.039** (0.543) | 0.579 | 0.380 |
| MCP-1 | **<0.0001 (0.002)** | 0.308 | **<0.0001 (<0.0001)** | **0.003 (0.018)** |
| MIP-1a | **0.0008 (0.007)** | **0.018** (0.506) | **0.0003 (0.001)** | **0.002 (0.018)** |
| MIP-1b | 0.089 | 0.219 | **0.0003 (0.001)** | **0.015** (0.085) |
| PDGF-bb | 0.736 | 0.463 | 0.058 | 0.434 |
| RANTES | 0.205 | 0.351 | **<0.0001 (0.0007)** | 0.058 |
| TNF-a | 0.167 | 0.505 | **0.002 (0.004)** | 0.089 |
| VEGF | 0.508 | 0.253 | **0.033 (0.048)** | 0.158 |

**Supplementary Table 5.** Joint fixed‑effects tests for longitudinal cytokine patterns

P‑values from joint F-tests were adjusted for multiple comparisons using the Benjamini–Hochberg false discovery rate procedure, applied separately within ART strata across all cytokines for each test (overall time effect and time × IUC interaction).
